# Life Events Extraction From Healthcare Notes for Veteran Acute Suicide Prediction

**DOI:** 10.1101/2025.02.03.25321612

**Authors:** Destinee Morrow, Rafael Zamora-Resendiz, Jean C. Beckham, Nathan A. Kimbrel, Benjamin H. McMahon, Silvia Crivelli

## Abstract

**Background/Aims:** Predictive models of suicide risk have focused on predictors extracted from structured data found in electronic health records (EHR), with limited consideration of predisposing life events (LE) expressed in unstructured clinical text such as housing instability and marital troubles. Additionally, there has been limited work in large-scale analysis of natural language processing (NLP) derived predictors for suicide risk and integration of extracted LE into longitudinal models of suicide risk. This study aims to expand upon previous research, demonstrating how high-performance computing (HPC) and machine learning technologies such as language models (LM) can be used to annotate and integrate 8 LE across all Veterans Health Administration (VHA) unstructured clinical text data with enriched performance metrics.

**Materials/Methods:** VHA-wide clinical text from January 2000 to January 2022 were pre-processed and analyzed using HPC. Data-driven lexicon curation was performed for each LE by scaling a nearest-neighbor search over a precomputed index with LM embeddings. Data parallelism was applied to a rule-based annotator to extract LE, followed by random forest for improved positive predictive value (PPV). NLP results were analyzed and then integrated and compared to a baseline statistical model predicting risk for a combined outcome (suicide death, suicide attempt and overdose).

**Results:** First-time LE mentions, with a PPV of 0.8 or higher, showed a temporal correlation to suicide-related events (SRE) (suicide ideation, attempt and/or death). A significant increase of LE occurrences was observed starting 2.5 months prior to an SRE. Predictive models integrating NLP-derived LE show an improved AUC of 0.81 vs. a 0.79 obtained with the baseline and novel patient identification of up to 57%.

**Discussion:** Our analysis shows that: 1) performance metrics, specifically PPV, improved significantly from previous work and outperform related works; 2) the mentions of LE in the unstructured data increase as time to a SRE approaches; 3) LE identified from the notes in the weeks prior to a SRE were not associated with administrative bias caused by outreach; and 4) LE improved the AUC of predictive models and identified novel patients at risk for suicide.

**Conclusion:** The resulting person-period longitudinal data demonstrated that NLP-derived LE served as acute predictors for suicide-related events. NLP integration into predictive models may help improve clinician decision support. Future work is necessary to better define these LE.

## 1 Background and Significance

Predictive models of suicide risk have traditionally focused on clinical predictors extracted from the structured data within electronic health records (EHR), such as demographics, diagnosis codes (i.e. International Classification of Diseases (ICD) codes), medications, and laboratory results.[1-4] Relatively few of these predictive models also consider negative or predisposing life events (LE) such as housing instability and social isolation, which are poorly represented in the structured EHR.[5] Systematic reviews conducted over the past five years demonstrated that an increasing number of studies are beginning to integrate social determinants of health (SDOH) into EHR predictive risk models.[6-11] Note that SDOH is a nomenclature used to either refer to social and community factors such as poverty level and crime rates, or to individual factors extracted from EHR such as LE. In this paper we use it for the latter.

SDOH examined in these review papers indicated a minimal to significant contribution to prediction, showcased by improved area under the curve (AUC). More than 60% of reviewed studies were conducted on sample sizes of less than 100,000 patients, which is a limitation because scaling of predictive models for health care systems presents additional statistical and logistic challenges. Less than 30% integrate the information annotated from unstructured clinical text into a predictive model (again reducing the potential impact of the NLP-derived LE). Overall, it is showcased that there are still many needed developments associated with SDOH, emphasizing the potential of ML and NLP.

Even though veterans remain at high risk of suicide, few studies have explored the vast and complex Veterans Health Administration (VHA) data to understand the effect of LE on veteran suicide-related risk.[12] Blosnich et al. used structured ICD codes to determine the prevalence of SDOH in VHA EHR and how SDOH were associated with suicide ideation and attempt.[13] Results showed that veterans with at least one SDOH had a 2.5 increase in the odds of suicidal ideation and a 3.3 increase in the odds of suicidal attempt; these numbers increased with a greater number of SDOH. Results only captured one years’ worth of patient data, which likely under-represents the VA patient population experiencing suicide-related events. Mitra et al. integrated NLP to annotate SDOH and evaluate suicide-related risk prediction using VHA data.[14] Their work highlighted the results of a fine-tuned RoBERTa model. Performance results did not show superior metrics when analyzing NLP even though a sophisticated LM was used. In our previous study, we focused on 9 LE that were identified by our VHA operational partners, as well as variables found in prior work to be strong predictors for accidental overdose and suicide death, including housing instability, job instability, food insecurity, criminal justice or troubles with the law, social connections specific to isolation and social connections specific to partner relationships, detoxification, military sexual trauma and access to lethal means.[5]

We demonstrated that a) an NLP pipeline could be used to extract these predisposing LE from unstructured data at a higher recall than structured VHA data, b) integration of these NLP factors with suicide risk models could improve performance, and c) this NLP pipeline could potentially improve targeting patients who have experienced LE for outreach by clinicians. We also showed that an important first step when extracting information from clinical notes is to understand the language being used. VHA healthcare prose and vocabulary found in unstructured clinical text can vary greatly across providers and subject matter domains, including jargon, misspellings, and abbreviations. This makes it difficult to construct a lexicon or dictionary that achieves high recall and precision. Identifying data-driven phrases with baseline statistical language models (LM) improves the total number of detectable mentions of LE but had moderate impact on positive predictive value (PPV).[5]

## 2 Objective

The objective of the current study was to evaluate whether our methodologies can be used to a) annotate LE with improved performance metrics (which can be used for streamlining future applications, b) identify acute suicide-related risk predictors without administrative bias and c) integrate LE into predictive models with improved accuracy and identification of novel patients at high risk for suicide-related events. Our HPC pipeline measures the prevalence of the NLP-derived LE across both a VHA-wide corpus and longitudinal patient timeline and it can be used to incorporate NLP-derived variables to existing and new predictive models and VHA-wide study designs.

## 3 Materials and Methods

### 3.1 Datasets

The analysis covered EHR data from the VHA CDW from over 23 million veterans, spanning from January 2000 to January 2022. Previous work explored the benefits of NLP-derived LE, specifically the performance improvement in recall when in comparison to structured data. Due to significant findings, this study focuses on how to improve upon LE annotation and positive predictive value (PPV) with more sophisticated NLP methodology, such as machine learning and state-of-the-art language models. Thus, we focus strictly on the available unstructured clinical text that is supplemented through physician notes, questionnaires, screeners, and more.

Building off the methodology in our previous study, we analyze the occurrence of predisposing LE across a VHA-wide patient cohort, requiring a vast amount of work involving the accessibility and integration of HPC to scale methods to the complete VHA cohort. We scaled LE annotations to a total of 2,289,783,296 ReportTexts generated from 1,783,395,072 care encounters for 23,550,293 patients (national PatientICN).

A dataset of 154,985 patients who had been flagged as high risk for suicide by suicide prevention coordinators were selected for additional analysis. For each patient, actions were recorded indicating one of four flagging response types: new, continued, reactivated, or deactivated. In this subset, new and reactivated flag events were observed between July 2012 and February 2022. Our aim is to investigate whether the accumulation of first-time LE precedes the new or reactivated flagging of a patient, thus indicating that these LE are not associated with administration bias. Further discussion can be found under 4.2. regarding methods to address administration bias.

### 3.2 Natural Language Processing Pipeline

We developed a pipeline, Figure 1, containing a suite of NLP tools, parallelized using message passing interface (MPI), that curates data-driven lexicons associated with 8

**Figure 1.**
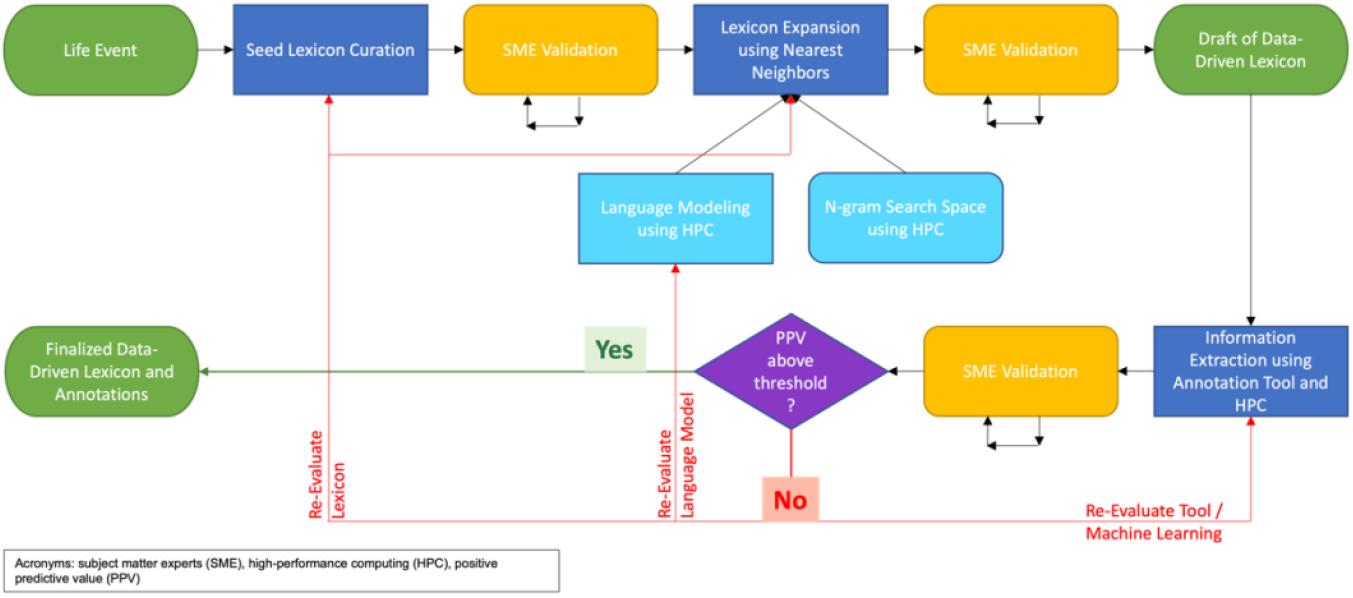
Natural Language Processing Pipeline An iterative natural language processing pipeline that incorporates the use of subject matter expects, language models, annotation tools and high-performance computing to curate data-driven lexicons and perform information extraction and annotation from unstructured clinical text for downstream analysis. N-gram refers to a sequence of a specified number of words in the unstructured clinical text.

LE, performs named-entity recognition (NER) over all VHA clinical text, and discriminates false-positive annotations. Detoxification was removed from this study design with reasonings explained in 5.1. Like previous work, this pipeline begins with selecting the LE of interest and curating an associated lexicon. A LM was used to search an VHA-wide n-gram space to create a data-driven lexicon for each of the LE. Next, an annotation tool was used to extract and annotate instances of the lexicon in unstructured clinical text. Subject matter experts (SME) performed iterative manual reviews of the lexicons and annotations until an acceptable inter-rater agreement of substantial (k >= 0.6) was achieved. Lastly, performance metrics were calculated and evaluated with an emphasis on PPV (a poor PPV of the model can create a significant clinical burden.) Additional steps such as re-evaluation of the lexicons, model training and or annotation tool, etc. may be necessary if PPV is not desirable. Each of the steps in this pipeline are described in more detail below and in the eMethods 1-3 of the supplemental material.

#### 3.2.2 Language Modeling

Many recent studies have found that newer, more sophisticated LM allow for improved information extraction and annotation and thus improved patient precision care. Si et al. compares frequently used LM such as Word2Vec, FastText, ELMo and BERT.[15] Performance results show an overall increased F1 score with BERT outperforming all models. Similarly, more recent studies, also using MIMIC data, assess how various architectures and frameworks perform when annotating SDOH.[16-18]

Our previous work evaluates the performance of simplistic Word2Vec to extrapolate data-driven concepts for clinical LE annotation and produced moderate performance results that could benefit from these more sophisticated LM.[5]

Preliminary analysis and comparison of various LM were performed and BioGPT-Large was selected for this study supported by compute-optimal ratios.[19] Note that these LM were not used for NER or classification due to hardware constraints, ease of scaling and pipeline at the time of analysis.

#### 3.2.3 Clinical Concept Annotation

We performed text analysis and information extraction and annotation using an NLP tool named Narrative Information Linear Extraction (NILE). NILE is a tree and rule-based tool that allows for NER in negation, location, modification, family history, and ignoring.[20] NILE processing can be abstracted into the following steps: 1) parse passages into tree data structures to search for target terms and modifiers, 2) match on target term and perform a suffix tree lookup for modifier terms, 3) assign a certainty value given combination of modifiers found. Other concept extraction tools like CLEVER differ in how the passages are parsed which changes the context window used to match on modifiers, as well as having different modifier classes and certainty rules.[21] We chose NILE due to the ease of scaling. Before running NILE over a set of clinical documents, the n-gram count vectorization is loaded to do an inverse lookup on the target n-grams. This reduces NILE processing to only those documents covered by the user defined lexicon. The total runtime of the NILE annotation depends on the size of the lexicon and how common the terms are.

#### 3.2.4 Temporal Analysis of LE

Understanding when these LE are occurring becomes pertinent when trying to predict the acute onset of suicide-related events. Because many of these events are captured within the unstructured clinical text there can be discrepancy between when the document is recorded and when the LE is occurring within the patient’s life. Thus, making it difficult to achieve high temporal precision and create a longitudinal timeline with these LE. As a first step to understanding the temporal alignment between structured date-times and NLP-derived LE, this study uses the structured data to construct a complete longitudinal patient timeline helping to establish first time mentions of LE. The use of structured date-times assumes that the LE is present and happening concurrent with the documentation. Our annotation tool, NILE uses a variety of temporal modifiers that adjust the assigned label according to the context surrounding the LE. Overall, NILE takes into consideration patient and family history of LE for temporal correlation. However, NILE annotations which are found to contain temporal modifiers, are not considered as first time mentions of LE. ML classification was used to refine and improve performance evaluation of temporality within annotations.

### 3.4 Machine Learning Refinement

The validation process indicated that NILE under-performed and did not meet our desired threshold of a PPV greater than 0.9. To help improve these metrics for downstream integration into predictive modeling and operations, we layered random forest (RF) classification to further predict absence or presence and temporality of these LE. Snippets were converted to bag of words (BoW) and in conjunction a 70/20/10 training, test and validation analysis was conducted for each of the LE. Performance comparison of NILE annotation and RF classification are reported in eTable 1 with inter-rater agreement reported in eTable 2 of the supplemental materials. ML provides improved performance metrics.

### 3.5 Statistical Analysis

We use a similar retrospective case control study design and logistic regression modeling as Dhaubhadel et al. to observe the additive effect of NLP-derived variables to structured variables on the discrimination of controls from patients reported as having died from suicide, having a suicide attempt or having an overdose.[22] Our models that integrate NLP-derived LE were evaluated and compared to baseline models integrating only structured variables. The outcome was stratified into patients with a recorded suicide death (SD), patients with a SD or suicide attempt (SA) and patients with a SD, SA or overdose (OD), referred to as ‘combined outcome’. Models were trained on a sampling of 528,653 patients which include 419,655 controls, 17,493 SD, 39,662 SA and 51,843 OD patients.

## 4 Results

This section analyzes the extraction and annotation of LE from the entire VHA CDW, a total of over 23 million patients, spanning 22 years (section 4.1) and from a subset of 154,985 patients flagged as high risk (section 4.2). It then analyzes the impact of adding LE to a predictive model with various outcomes (section 4.3).

### 4.1 Longitudinal Life Events

Figure 2 shows the accumulation of new LE over time. A trend showing a spike in new first-time LE mentions can be seen starting 2.5 months prior to their suicide-related event. Log-scaled axis was used to show the concentration of multiple LE over narrowing windows of time before the suicide-related events. Interestingly, more than 10,000 patients recorded a mention for at least 5 unique LE the week prior their first time suicide ideation event. Along first-time suicide ideation and suicide attempt, 10% and 6% patients had records indicating at least 1 new LE the week prior to diagnosis respectively. A similar plot showcasing the weeks prior to suicide death and first-time renal failure for objective comparison of prevalence can be found in eFigure 1 of the supplemental materials.

**Figure 2.**
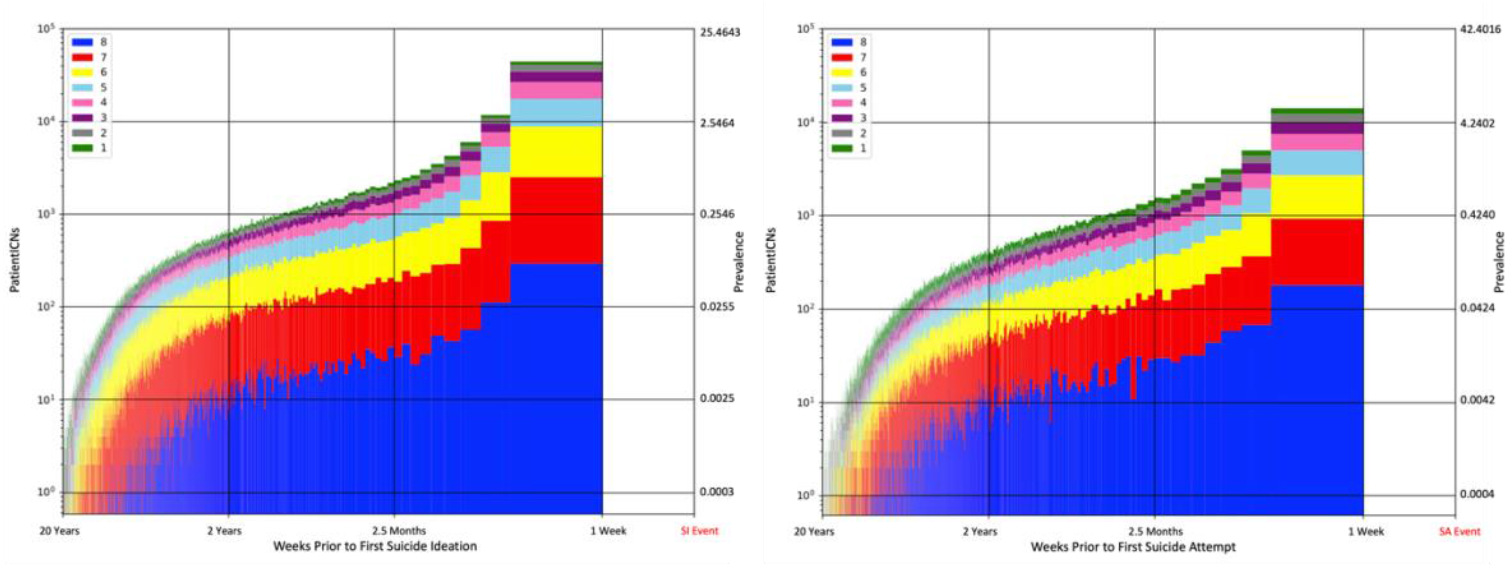
Longitudinal Life Events Number of patients per week receiving their Nth unique LE, N=1,..8. The first occurrence of positive LE mentions was aligned to the date of the respective first-time suicide-related event; suicide ideation (SI) (left), suicide attempt (SA) (right). Prevalence is reported as a percentage of the number of patients with suicide ideation (n=392,706) and suicide attempt (n=235,840) respectively.

### 4.2 Administrative Bias Consideration

While the VHA-wide cohort shows a strong temporal correlation between the accumulation of unique LE and proximity to a first-time ideation or attempt event, cross-validation was done to ensure the accuracy of the LE annotation and discard potential sources of administrative bias found near the suicide event. In particular, mental health and suicide prevention services are known to survey for risk factors as part of triaging patients with high risk of suicidality. To determine the performance of our LE annotation with respect to detecting the onset of suicide ideation and attempts, we conducted a sub-analysis of mentions along the subset of patients who have been flagged as high risk for suicide by a suicide prevention coordinator. This allows for a clearer, non-biased, interpretation of the prevalence of LE with respect to suicide-related events. Further explanation and results of this analysis can be found in eMethods 4 and eFigure 2 of the supplemental materials.

### 4.3 Impact on Predictive Modeling

Our methods highlight which of these LE and how many LE contribute most to risk. Figure 3 shows that LE have a positive coefficient or predictive impact on discriminating controls from patients with a combined outcome. All the LE, excluding food insecurity, as well as, having more than 1 co-occurring LE (eFigure 3 of the supplemental materials) are predictive for a combined outcome. Coefficients for individual and co-occurring LE for the SD and SD or SA outcome are found in eFigures 4-5 of the supplemental materials. Table 1 showcases that those models which integrate NLP-derived LE slightly outperform baseline models that integrate only structured variables. Top 1 and 0.1 percent of the high-risk patients identified from the logistic regression models are reported in Table 2 and demonstrate that the model including NLP-derived LE identifies more than 40 percent of patients not identified in the baseline model. This table also showcases that on average more than 46 and 55 percent of the top 1 and 0.1 respectively are experiencing LE, highlighting the importance of including these LE in predictive models.

**Figure 3.**
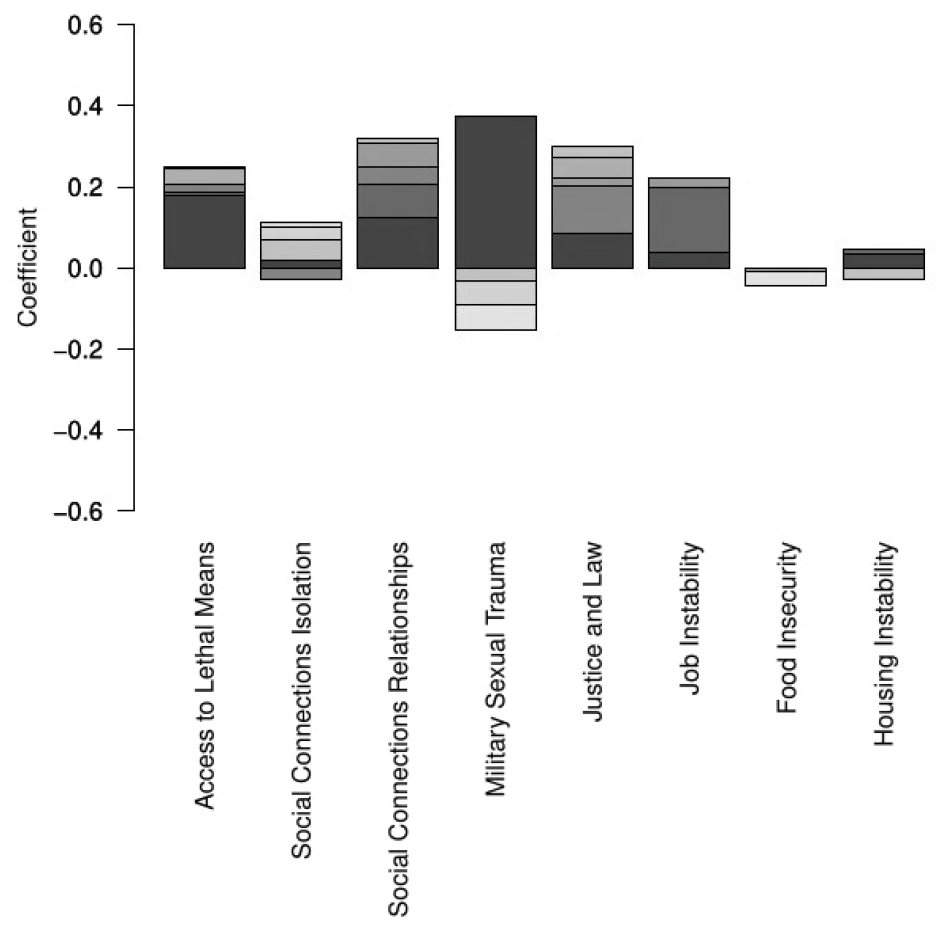
LE Coefficients for the Combined Outcome Adjusted logistic regression coefficients for each of the LE. Each bar represents cumulative impact on predicted risk of the combined outcome (suicide death, suicide attempt and or overdose) for presence of the variable along 8 time bins. Shade of sections within the bars indicates proximity to outcome date (darker indicating closer proximity than lighter).

**Table 1.** Model Performance.

| Model | Outcome | AUC (95% CI +/-) |
| --- | --- | --- |
| Baseline | SD | 0.767 (0.0035) |
| Baseline + LE | SD | **0.771** (0.0035) |
| Baseline | SD/SA | 0.786 (0.0021) |
| Baseline + LE | SD/SA | **0.787** (0.0021) |
| Baseline | Combined | 0.790 (0.0028) |
| Baseline + LE | Combined | **0.806** (0.0015) |

Model performance stratified by variables included and predicted outcome. Models include those that incorporate NLP-derived LE as well as structured variables (Baseline + LE) and just structured variables (Baseline). Predicted outcomes include suicide death (SD), suicide death or suicide attempt (SD/SA) and combined outcome of suicide death, suicide attempt or overdose.

**Table 2.** Top 1 and 0.1% Suicide-Related Risk.

|  | Top 1% | Top 1% |  | Top 0.1% | Top 0.1% |  |
| --- | --- | --- | --- | --- | --- | --- |
|  | Baseline | Baseline + LE | Novel | Baseline | Baseline + LE | Novel |
| Access to Lethal Means | 1949 | 2691 | 991 | 239 | 487 | 273 |
| Social Connections Isolation | 1985 | 2713 | 998 | 241 | 492 | 295 |
| Social Connections Relationships | 1963 | 2732 | 1021 | 233 | 490 | 270 |
| Military Sexual Trauma | 413 | 540 | 193 | 55 | 104 | 60 |
| Justice and Law | 1890 | 2596 | 957 | 238 | 499 | 279 |
| Job Instability | 1945 | 2654 | 1002 | 238 | 506 | 281 |
| Food Insecurity | 1269 | 1668 | 560 | 178 | 351 | 197 |
| Housing Instability | 2028 | 2793 | 1042 | 241 | 502 | 301 |
| Total | 3,510 | 4,971 | 2,018<br>(40.60%) | 391 | 783 | 444<br>(56.70%) |

Number of unique patients identified in the top 1 and 0.1 percent of high risk, from a test set of 377,691 patients, by the baseline and baseline with incorporated NLP-derived LE logistic regression models trained on the combined outcome. Results are stratified by which LE these patients are experiencing based on the NLP model. Number of novel patients, those that were not identified in top 1 and 0.1 percent by the baseline logistic regression, are also reported.

### 4.4 Additional Analyses

A breakdown of the number of unique patients, visits, and documents identified by our NLP pipeline per LE and cohort is reported in eTable 3 of the supplemental materials.

Renal failure is again used for objective comparison of LE prevalence. Various quality assurance analyses were performed for explainability of the statistical modeling.

Bivariate logistic regression per LE reported in eTable 4 of the supplemental materials shows that all NLP-derived LE have significant impact with odds ratios ranging between 8.29 (MST) to 15.78 (A2M). Adjusted logistic regression coefficients for all the structured variables included in the model were analyzed for comparison to previously reported results by Dhaubhadel et al. and are reported in eFigure 6 of the supplemental materials.[22]

## 5 Discussion

As hoped, our NLP pipeline, which uses both LM and ML, was able to meet multiple performance metrics at scale. Two of the LE reached the desired threshold of 0.9 PPV, with 4 closely trailing behind. Although not a 1-to-1 comparison, many of our similar LE outperformed the exact and relaxed performance metrics reported by Mitra et al., the most relevant related work, for their SDOH.[14] Additionally, both suicide ideation and attempt patients both showed an increase in LE as time approaches the suicide-related event. LE can be seen as distant as 20 years prior and as close as 1 week before. They seemed to have a compound effect on these suicide-related events as an increasing number of patients appeared to experience new LE less than 3 months before the occurrence of the suicide-related event.

Additionally, our results also suggested that mentions are independent from administrative bias introduced by clinician outreach assuming the patients had not received outreach by suicide prevention or mental health departments prior to the high-risk flag events. Our findings further suggest that the accumulation of first-time LE may be a potential early indicator of future suicide-related risk.

The inclusion of NLP-derived LE into predictive models used to discriminate controls from any patient outcome (suicide death, suicide attempt or overdose) showcased slightly improved AUC. Logistic regression coefficients varied depending on the encoding and patient outcome. Regardless, for those models including suicide attempt and or overdose, it was clear that the LE were predictive and added value to the models. Models discriminating controls from suicide deaths demonstrated more variability which may be attributed to the low statistical power discussed in eMethods 5 of the supplemental material. Both the longitudinal and logistic regression analysis found that 3 or more LE were acute risk predictors of suicide-related events.

Overall, scaling of our NLP pipeline allowed us to run weak labeling over the entire CDW to annotate 8 LE. Our analysis shows that: 1) performance metrics, specifically PPV, improved significantly from previous work and outperform related works; 2) the mentions of LE in the unstructured data increase as time to a suicide-related event approaches; 3) a combination of up to 8 LE were identified from the notes in the week prior to a suicide ideation or attempt event; 4) LE identified from the notes in the weeks prior to a suicide ideation or attempt event were not associated with administrative bias caused by outreach; 5) a combination of 3 or more LE are acute risk predictors of suicide-related events; and 6) LE improved AUC of predictive models and identified novel patients at risk for suicide.

### 5.1 Limitations and Future Work

Our study also had some limitations that should be considered while interpreting these findings.

Lituiev et al. highlighted similar limitations that are associated with analyzing SDOH using similar NLP methods.[23] Examples of these limitations include false positives due to varying difficulty of extraction per LE. False positives are commonly seen with annotation tools’ inability to decipher semantics and template text, which abounds in the clinical notes. Varying performance between LE may also be attributed to a fluid, non-static definition of the LE across the 22-year analysis, including changes in clinical screenings and questionnaires that are difficult to address. Lastly, to accurately annotate the fine-grained temporal component of all LE mentions (e.g. clarification of a LE happening 2 weeks vs. 2 years in the past) may require a more sophisticated ML or large LM approach. Future work will move in the direction of state-of-the-art large LM to help combat these limitations emphasizing the need to reach our desired threshold of a PPV greater than 0.9 for all LE. However, a recent study by Shortreed et al. may suggest detailed temporal annotation is unnecessary, highlighting minimal performance gains to Chen et al. who uses similar methodologies to this study.[24-25]

Our study does not take into consideration the risk assessment bias potentially introduced by annual screenings for LE such as access to lethal means, food insecurity and housing instability that is performed on all patients regardless of if they have or have not been flagged as high risk for suicide.

Our study did not compare or adjust LE definitions to other related literature which makes comparisons and reflections between related literature difficult. Lexicons used in this study are reported in eTable 5 of the supplemental materials to facilitate others’ contributions towards the creation of standards. Expansion of LE and addition of related concepts will be considered in future work to help improve sensitivity and reproducibility of these suicide-related events. Our study used a limited number of samples for training, testing and validation in comparison to the size of analysis.

Previous work included detoxification as a LE, which was removed from this analysis. This was due to multiple reasons including raters could not reach the necessary moderate agreement for this LE. Future work is needed to revisit and establish a better definition for this LE.

Finally, NLP may be capturing redundant information already considered by the structured variables. This may be in part attributed to the large use of screeners, questionnaires and templates that are documented in the unstructured clinical text. Additional pre-processing of the notes and or more sophisticated LM may be required to evaluate and reduce redundancy.

## 6 Conclusion

LE can be captured longitudinally at scale using our NLP pipeline with performance metrics comparable to related works. Our results continually showcase the importance of integrating these LE into downstream applications such as predictive models that typically exclude such information.

### 6.1 Clinical and Research Implications

This study has various important research and clinical implications. To our knowledge this is the first analysis of these LE across all available VHA data and the first to include these LE into a predictive model and assess its performance improvements. At this magnitude, there is now a baseline prevalence established for each of these LE, including the prevalence of patients with suicide-related events experiencing these LE. Our data structure, pipeline and documentation ensure ease of use and indexing that is available for future VA researchers. Our pipeline can be applicable to other datasets and use cases. Clinician decision support and outreach may see improvement with the identification of patients experiencing LE which serve as acute predictors of suicide-related events. Clinicians may wish to implement earlier outreach based on when patients start experiencing these LE, potentially lowering the rate of suicide-related events.

## Data Availability

All data is not available.

## 6.2 Acknowledgments

This research is based on data from the Million Veteran Program, Office of Research and Development, Veterans Health Administration, and was supported by MVP062. This publication does not represent the views of the Department of Veteran Affairs or the United States Government. Dr. Beckham was supported by a Senior Research Career Scientist Award from CSRD (lK6BX003777). Dr. Kimbrel was supported by a Research Career Scientist Award from VA BLRD (IK6BX006523).

