## Supplemental Materials for "Life Events Extraction From Healthcare Notes for Veteran Acute Suicide Prediction"

Supplemental Online Content


eMethods 1. Lexicon Curation
eMethods 2. Validation
eMethods 3. Inter-rater Agreement
eTable 1. Annotation Performance Metrics
eTable 2. Inter-rater Agreement
eFigure 1. Longitudinal Life Events
eMethods 4. Administrative Bias Consideration
eFigure 2. Longitudinal Life Events Administrative Bias Consideration
eFigure 3. Co-Occurring LE Coefficients for the Combined Outcome
eFigure 4. Life Events Coefficients for the Suicide Death or Suicide Attempt Outcome
eFigure 5. Co-Occurring Life Events Coefficients for the Suicide Death or Suicide Attempt Outcome
eTable 3. Life Events Prevalence
eTable 4. Life Events Odds Ratios
eFigure 6. Structured Variable Coefficients
eMethods 5. Suicide Death Cohort Exploration
eTable 5. Lexicon
References

### eMethods 1. Lexicon Curation

In this study, we use the same seed terms that we used in our previous work but change the language modeling and statistical methodology for lexicon expansion.[1]

In conjunction with language model (LM) embeddings (3.2.2), a search space was generated that would allow for the inclusion of all possible combinations of life event (LE) notation. Using message passing interface (MPI), the framework was first used to gather unique n-grams (unigram, bigram, and trigram), a sequence of a specified number of words in the unstructured clinical text, across the Veterans Health Administration VHA-wide corpus using sklearn's CountVectorizer. Additionally, sparse matrices of the resulting count vectorizations were stored per individual partition. These sparse matrices can be used as bag-of-words features for machine learning or as an inverse lookup for documents containing a specified set of n-grams. Each n-gram was hashed using SHA224 to uniquely identify terms across the VHA corpus. A total of 164,870,027 n-grams, occurring over 10 or more unique documents, were indexed. Out of vocabulary (OOV) searching proves to be useful as it reveals nearest neighbors that normally would not be found in the training corpus. This helps to deal with the complex syntax, formatting and terminology commonly found within clinical text. Thus, this method allows for all possible data-driven nearest neighbors to be considered.

A subset of 19,888,205 n-grams were used to perform lexicon expansion for the 8 LE. This set of n-grams was used as the search space for nearest neighbor expansion w.r.t. the seed lexicons previously constructed by subject matter experts (SME). The top 1000 nearest neighbor results for each LE were reviewed and included if applicable.

### eMethods 2. Validation

The validation process consisted of 2 independent raters reviewing the annotated LE mentions leading up to the suicide-related events on a subset of patients marking the correct assessment of certainty (presence or absence; patient is the experiencer) and temporality (currently happening or historical). We measure the recall, positive predictive value (PPV) and F1-score.

Related work has shown inconsistencies regarding the acceptable number of documents to use during validation, or when creating a gold standard set. Although we believe methods developed by Juckett et al. justify a sound curation of a gold standard set, these techniques are not plausible for the size of this analysis.[2] Thus, we sampled a set of snippets from all available annotations for our validation that we believed would be feasible and justify our methods. Snippets include the LE of interest and a pre-determined context window. The set contains 500 randomly sampled patients with a ratio of 1 to 4 for cases to controls, stratified by lexicon terminology, allowing us to observe the overall performance of annotation per LE.

eMethods 3. Inter-rater Agreement

Cohen's Kappa was introduced to account for the probability of chance agreement between raters. This statistic represents the extent to which the labels are accurate representations of the observed variables.[3] It is common practice to use this statistic when two raters are involved and assessing all concepts. k = (Po-Pe)/(1-Pe) Where Po = observed agreement among raters and Pe = probability of chance agreement. The k statistic ranges from [0, 1] where 0 is agreement equivalent to chance, [0.1, 0.2) is slight agreement, [0.2, 0.4) is fair agreement, [0.4, 0.6) is moderate agreement, [0.6, 0.8) is substantial agreement, [0.8, 1.0) is near perfect agreement and 1 indicates perfect agreement between raters. Inter-rater agreement between reviewers across LE can be found eTable 2.

eTable 1. Annotation Performance Metrics

| LE | Method | Recall | PPV | F1 Score |
|---|---|---|---|---|
| Access to Lethal Means | NLP<br>ML | 0.81<br>0.71 | 0.40<br>**0.68** | 0.54<br>**0.65** |
| Social Connections Isolation | NLP<br>ML | 0.91<br>0.76 | 0.55<br>**0.76** | 0.68<br>**0.76** |
| Social Connections Relationships | NLP<br>ML | 0.90<br>0.85 | 0.52<br>**0.85** | 0.66<br>**0.85** |
| Military Sexual Trauma | NLP<br>ML | 0.95<br>0.90 | 0.57<br>**0.92** | 0.71<br>**0.90** |
| Justice and Law | NLP<br>ML | 0.90<br>0.79 | 0.52<br>**0.80** | 0.66<br>**0.78** |
| Job Instability | NLP<br>ML | 0.86<br>0.84 | 0.48<br>**0.86** | 0.61<br>**0.83** |
| Food Insecurity | NLP<br>ML | 0.94<br>0.90 | 0.71<br>**0.91** | 0.81<br>**0.91** |
| Housing Instability | NLP<br>ML | 0.91<br>0.86 | 0.62<br>**0.86** | 0.74<br>**0.85** |

Performance metrics of natural language processing (NLP) (NILE annotation) and machine learning (ML) (Random Forest Classifier) methods. Recall, positive predictive value (PPV) and F1 score are reported.

eTable 2. Inter-Rater Agreement

| LE | Samples | Cohen’s Kappa |
|---|---|---|
| Access to Lethal Means | 500 | 0.69 |
| Social Connections Isolation | 500 | 0.99 |
| Social Connections Relationships | 500 | 0.98 |
| Military Sexual Trauma | 500 | 0.84 |
| Justice and Law | 500 | 0.75 |
| Job Instability | 500 | 0.65 |
| Food Insecurity | 500 | 0.79 |
| Housing Instability | 500 | 0.71 |

Validation was conducted between two independent reviewers and agreement between labels was measured using Cohen’s Kappa.

eFigure 1. Longitudinal Life Events

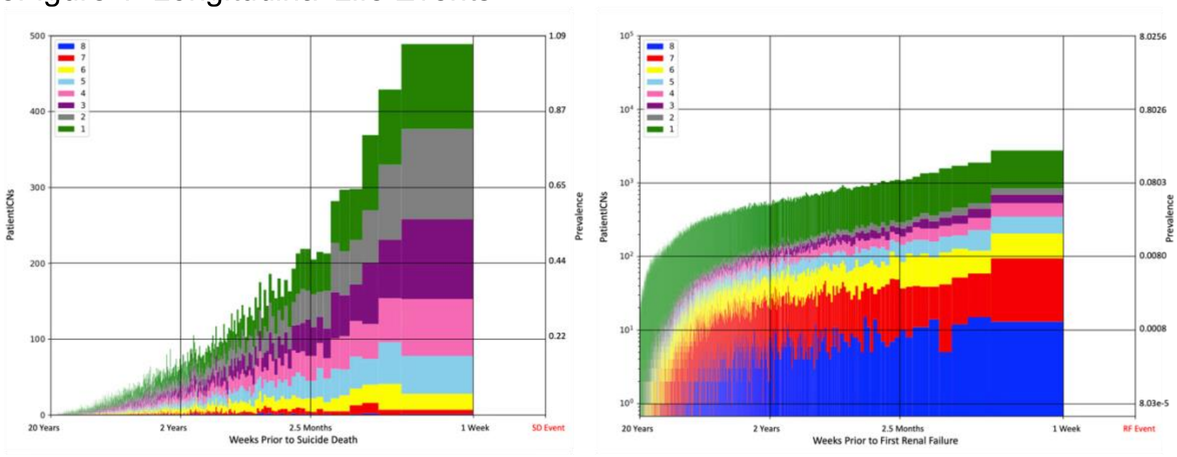


Number of patients per week receiving their Nth unique LE, N=1,..8. The first occurrence of positive LE mentions was aligned to the date of the respective suicide death event (left) and first-time renal failure event (right). Prevalence is reported as a percentage of the number of patients with a suicide death (n=45,899) and renal failure (n=1,246,002) respectively.

eMethods 4. Administrative Bias Consideration

We have cross-validated the patients seen in Figure 2 to ensure that they have no data related to suicide risk assessment prior to the new flag or reactivated flag date. The identification of first-time LE mentions prior to the recorded date of flagging would indicate that the extracted mentions are less likely to be from clinical text generated during the triaging of suicidal patients. Of the 154,985 patients who have been flagged as high risk for suicide, 96,088 and 54,001 were diagnosed with suicide ideation and attempt, respectively, and had LE mentions prior to the suicide-related diagnosis. Approximately 85% of the suicide attempt patients were diagnosed with ideation as well. It should be noted that not all patients that receive a high-risk flag will also receive a suicide ideation or suicide attempt diagnosis code.

eFigure 2. Longitudinal Life Events Administrative Bias Consideration

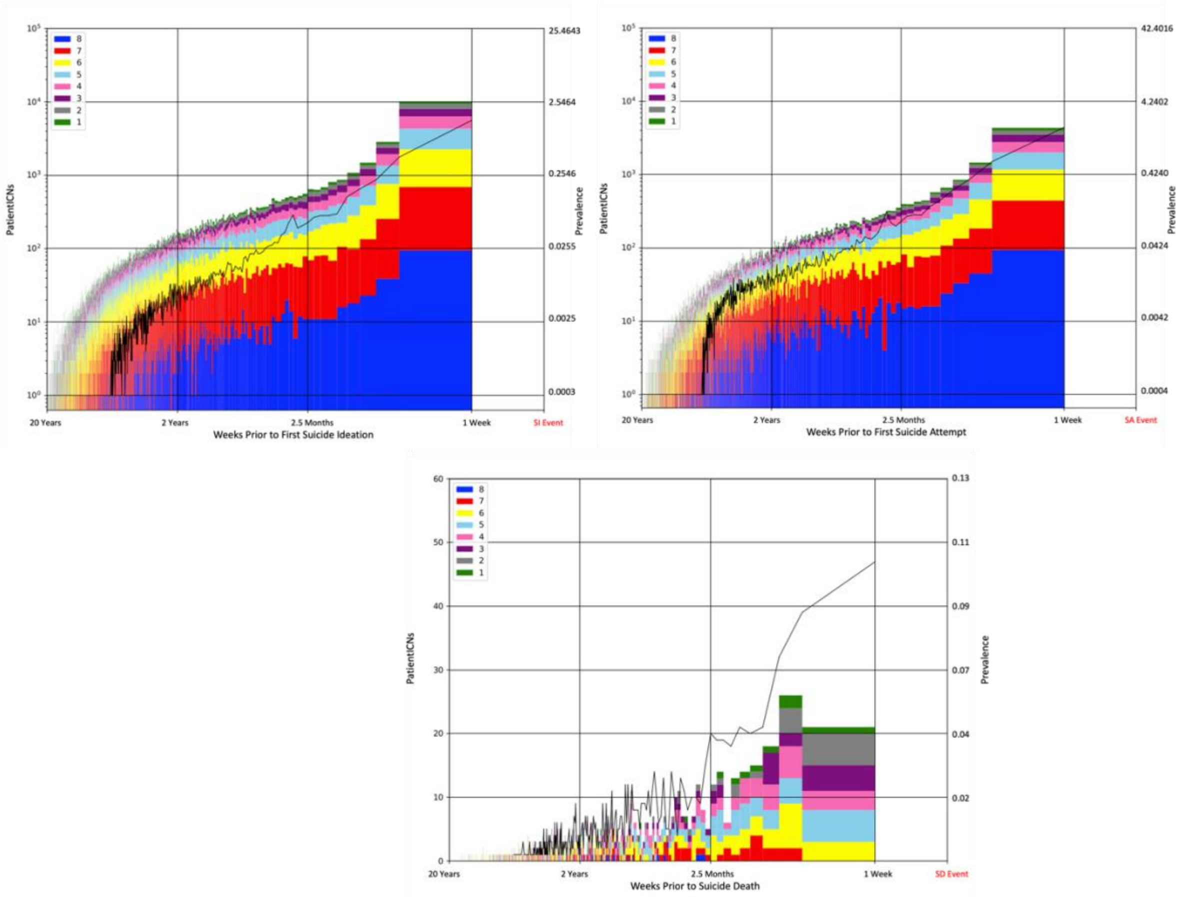


Number of patients per week receiving their Nth unique LE, N=1,..8. The first occurrence of positive LE mentions was aligned to the date of the respective first-time suicide-related event; suicide ideation (SI) (left), suicide attempt (SA) (right) and suicide death (SD) (bottom). Prevalence is reported as a percentage of the number of patients with suicide ideation (n=392,706), suicide attempt (n=235,840) and suicide death (n=45,899) respectively. The black line represents the number of patients being flagged per week.

eFigure 3. Co-Occurring LE Coefficients for the Combined Outcome

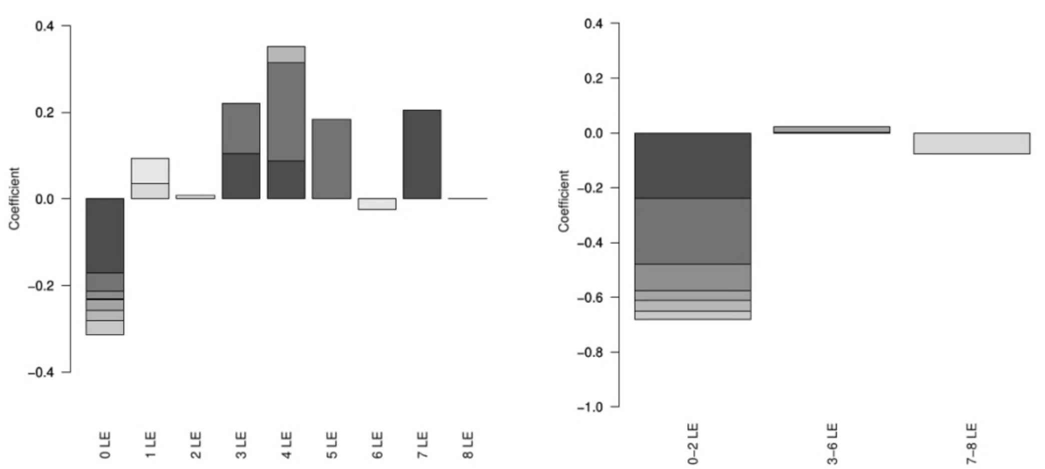


Adjusted logistic regression coefficients for co-occurring number of LE. Each bar represents cumulative impact on predicted risk of the combined outcome (suicide death, suicide attempt or overdose) for presence of the variable along 8 time bins. Shade of sections within the bars indicates proximity to outcome date (darker indicating closer proximity than lighter).

eFigure 4. LE Coefficients for the Suicide Death or Suicide Attempt Outcome

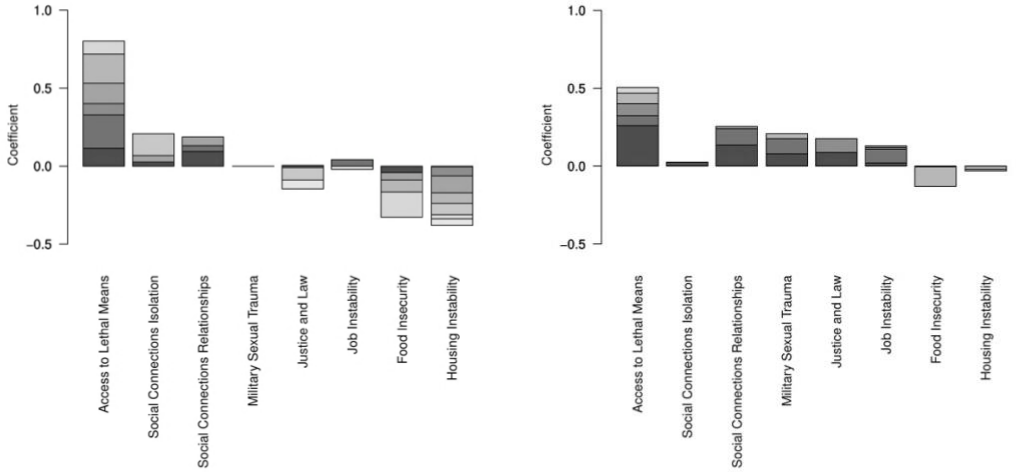


Adjusted logistic regression coefficients for each of the LE. Each bar represents cumulative impact on predicted risk of suicide death (left) and suicide death or suicide attempt (right) outcome for presence of the variable along 8 time bins. Shade of sections within the bars indicates proximity to outcome date (darker indicating closer proximity than lighter).

eFigure 5. Co-Occurring LE Coefficients for the Suicide Death or Suicide Attempt Outcome

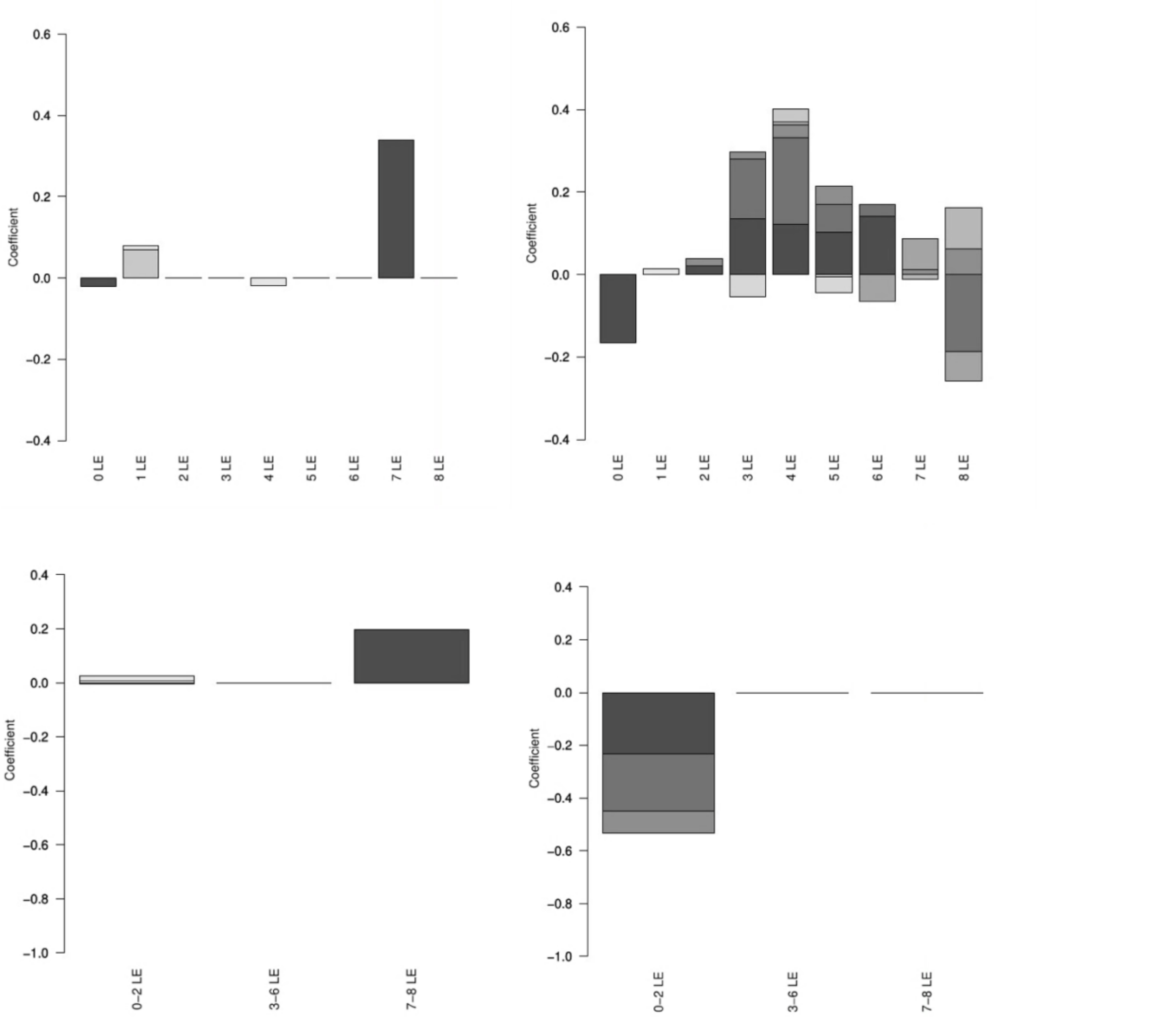


Adjusted logistic regression coefficients for co-occurring number of LE. Each bar represents cumulative impact on predicted risk of suicide death (left) and suicide death or suicide attempt (right) outcome for presence of the variable along 8 time bins. Shade of sections within the bars indicates proximity to outcome date (darker indicating closer proximity than lighter).

eTable 3. LE Prevalence

| Life Event | Cohort | PatientICNs | VisitSIDs | TIUDocuments |
|---|---|---|---|---|
| Access to Lethal Means | Overall (n=23,550,293) | 1,460,225 (6.20%) | 3,914,910 | 4,300,737 |
| | Suicide Ideation (n=392,706) | 219,875 (55.99%) | 108,574 | 1,237,850 |
| | Suicide Attempt (n=235,840) | 108,574 (46.04%) | 536,495 | 633,252 |
| | Suicide Death (n=45,899) | 7,811 (17.02%) | 20,387 | 24,883 |
| | Renal Failure (n=1,246,002) | 216,520 (17.38%) | 593,857 | 651,826 |
| Social Connections Isolation | Overall | 4,596,934 (19.52%) | 22,489,990 | 25,905,937 |
| | Suicide Ideation | 296,701 (75.55%) | 3,803,793 | 4,740,395 |
| | Suicide Attempt | 168,629 (71.50%) | 2,145,341 | 2,707,298 |
| | Suicide Death | 15,773 (34.36%) | 72,966 | 91,795 |
| | Renal Failure | 674,432 (54.13%) | 4,551,371 | 5,290,158 |
| Social Connections Relationships | Overall | 3,785,476 (16.07%) | 19,687,246 | 21,754,526 |
| | Suicide Ideation | 290,587 (73.98%) | 4,290,088 | 5,070,628 |
| | Suicide Attempt | 159,371 (67.58%) | 2,332,326 | 2,789,076 |
| | Suicide Death | 16,609 (36.19%) | 87,155 | 107,706 |
| | Renal Failure | 515,343 (41.36%) | 3,011,850 | 3,336,810 |
| Military Sexual Trauma | Overall | 482,072 (2.05%) | 3,620,222 | 3,864,637 |
| | Suicide Ideation | 75,528 (19.23%) | 1,130,916 | 1,254,751 |
| | Suicide Attempt | 38,834 (16.47%) | 628,151 | 700,455 |
| | Suicide Death | 1,110 (2.42%) | 7,425 | 8,244 |
| | Renal Failure | 50,997 (4.09%) | 400,939 | 430,371 |
| Justice and Law | Overall | 2,060,526 (8.75%) | 10,543,928 | 11,927,572 |
| | Suicide Ideation | 251,130 (63.95%) | 3,350,007 | 3,984,596 |

| | | | | |
|---|---|---|---|---|
| | Suicide Attempt | 129,923 (55.09%) | 1,837,113 | 2,211,581 |
| | Suicide Death | 10,263 (22.36%) | 52,189 | 63,581 |
| | Renal Failure | 296,280 (23.78%) | 1,375,168 | 1,565,336 |
| Job Instability | Overall | 2,944,009 (12.50%) | 14,654,822 | 16,564,075 |
| | Suicide Ideation | 274,568 (69.92%) | 3,894,186 | 4,760,585 |
| | Suicide Attempt | 145,476 (61.68%) | 2,051,254 | 2,544,236 |
| | Suicide Death | 13,342 (29.07%) | 66,129 | 81,770 |
| | Renal Failure | 381,388 (30.61%) | 1,943,824 | 2,198,094 |
| Food Insecurity | Overall | 916,234 (3.89%) | 2,742,711 | 2,855,199 |
| | Suicide Ideation | 124,114 (31.60%) | 568,958 | 592,713 |
| | Suicide Attempt | 64,390 (27.30%) | 294,010 | 307,170 |
| | Suicide Death | 2,619 (5.71%) | 5,678 | 5,997 |
| | Renal Failure | 165,670 (13.30%) | 545,539 | 570,279 |
| Housing Instability | Overall | 2,785,857 (11.83%) | 48,922,198 | 59,923,669 |
| | Suicide Ideation | 272,829 (69.47%) | 17,955,396 | 23,066,387 |
| | Suicide Attempt | 144,013 (61.06%) | 9,540,846 | 12,442,289 |
| | Suicide Death | 11,085 (24.15%) | 145,985 | 199,133 |
| | Renal Failure | 388,051 (31.14%) | 7,066,183 | 8,682,243 |
| Total | | 7,036,469 | 102,886,258 | 124,131,808 |

The number of unique patients captured along the NLP LE annotations, containing at least 1 positive LE mention, as well as the unique number of visits and clinical documents.

eTable 4. Life Events Odds Ratios

| LE | Coefficients | OR |
|---|---|---|
| Access to Lethal Means | 5.806 | 15.782 |
| Social Connections Isolation | 4.210 | 11.443 |
| Social Connections Relationships | 3.819 | 10.380 |
| Military Sexual Trauma | 2.839 | 7.716 |
| Justice and Law | 4.572 | 12.429 |
| Job Instability | 4.117 | 11.190 |
| Food Insecurity | 5.090 | 13.836 |
| Housing Instability | 3.050 | 8.290 |

Logistic regression bivariate analysis of each LE and the combined outcome (suicide death, suicide attempt or overdose). Their respective logistic regression coefficient and odds ratio (OR) is reported.

eFigure 6. Structured Variable Coefficients

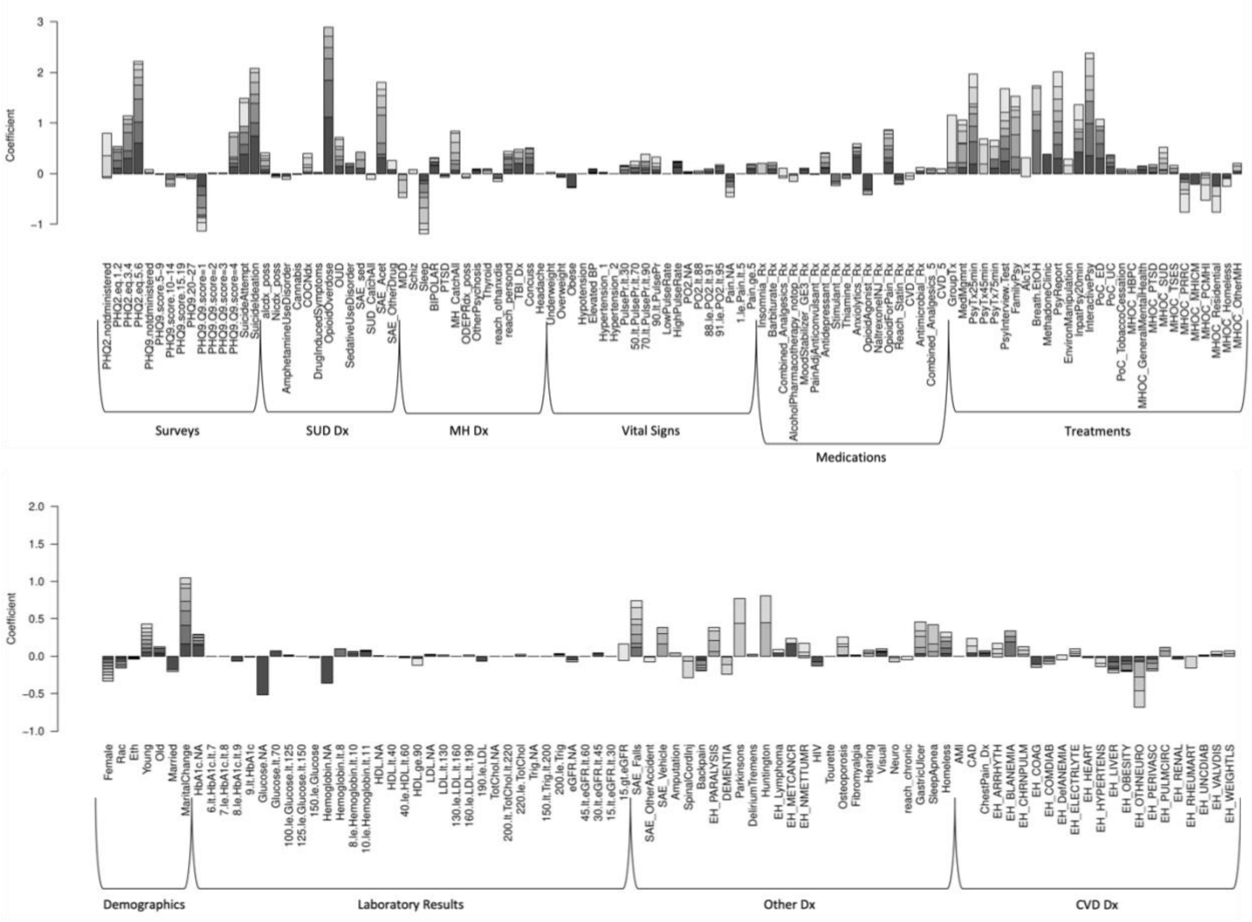


Adjusted logistic regression coefficients for each of the structured variables. Each bar represents cumulative impact on predicted risk of the combined outcome (suicide death, suicide attempt or overdose) for presence of the variable along 8 time bins. Shade of sections within the bars indicates proximity to outcome date (darker indicating closer proximity than lighter). Structured variable definitions are reported in the Dhaubhadel et al. manuscript.[4]

eMethods 5. Suicide Death Cohort Exploration
Additional exploration of the NLP results also shows that there are 17,011 suicide deaths with no clinical text records indicating at least one of the 8 LE categories explored in the study demonstrated in Table 3. From this set of patients, 2,376 patients did not have TIUDocuments across the 22-year dataset. Additionally, only 4,586 patients had documents containing the regex r'suicide|suicidal'. Total PatientICNs, VisitSIDs and TIUDocumentSIDs for this regex is 7,205,537, 89,170,247 and 100,737,464 respectively. This leaves a total of 10,049 suicide deaths not covered by the current configuration of LE or suicide mention extractors. We ranked terms from this outlier subset using Term Frequency - Inverse Document Frequency (TF-IDF) to look for terms that may characterize this subset. The following are the terms and their respective prevalence:
'pain' – 7,864 patients (78\%); 'cancer|carcinoma' – 5,511 patients (55\%); 'chronic' – 4,545 patients (45\%); 'hepatitis' – 4,375 patients (44\%); 'ptsd' – 3,092 patients (31\%); Union of terms – 8,370 patients (83\%).
1,679 patients did not have these additional target terms, and their notes were sampled for reading. After reading through the set of notes, we corroborated that they correspond to suicide death with low quantity of textual data and/or deaths which occurred closer to January 2000 when clinical text recording was less prevalent.

eTable 5. Lexicon

| Life Event | Length | Concepts |
|---|---|---|
| Access to Lethal Means | 78 | agun, airsoft, antifreeze, asphyxiate, asphyxiating, asphyxiation, axe, bayonet, boxcutter, bullets, carbine, chainsaw, crossbow, crowbar, cyanide, electrocute, fire arms, firearems, firearm, firearmes, firearms, glock, gun, gunlock, guns, gunsafe, gunshop, handgun, handguns, hangun, hatchet, jump off bridge, knife, knifed, knifes, knive, knives, knofe, machete, machetes, machette, machetti, machinegun, magnum, monoxide, musket, muzzle, muzzleloader, noose, pistal, pistol, pistols, pocketknife, purchase ammunition, revolver, revolvers, riffles, rifle, rifles, rope, roulette, securefirearms, semiautomatic, shotgun, shotguns, submachine, suffocate, switchblade, sword, weapoms, weapon, weaponry, weapons, weopon, weopons, wepaons, wepon, wepons |
| Social Connections Isolation | 73 | abandoment, abandonement, abandonment, abandonments, agoraphobic, alienated, alienating, alienation, alientated, aloneness, aloofness, avoidant, despondent, difficulty establishing relationships, difficulty maintaining relationships, distanced, distancing, estranged, estrangement, estrangment, family conflict, family disconnect, family discord, family distant, family isolation, family support lacking, fears people, feeling lonely, home isolation, inadequate social skills, inadequate social support, interpersonal conflict, interpersonal stressor, irritability social, isolation withdrawal, isolative, lacks social support, limited social interaction, limited social support, lives alone, living alone, living situation alone, loneliness, lonely, lonelyness, loner, loners, lonesome, lonley, lonlieness, lonliness, lonly, no social support, ostracized, outcast, patient live alone, patient resides alone, poor social support, |

| | | |
|---|---|---|
| | | problems social, relationship difficulties, relationships broken, social exclusion, social interaction impairment, social isolation, social outcast, social paranoia isoation, social rejection, social support lack, social support lacks, social support lonely, social withdrawal, socially isolated, solitude |
| Social Connections Relationships | 94 | abuse and domestic, abuse domestic, abuse neglect domestic, affair, assault molestation domestic, behavior interpersonal violence, current relationship issues, devorce, devorced, divirce, divirced, divoce, divoced, divorce, divorced, divorcedx, divorcee, divorces, divorcing, divorded, divore, divoreced, divored, divorice, divoriced, divorse, divorsed, divroced, domestic abuse, domestic assault, domestic partner violence, domestic violence, exhusband, exwife, exwifes, exwives, family conflict, family violence, fights family violence, financial strain marital, husband left home, infedelity, infidelities, infidelity, interpersonal violence or, intimate partner abuse, intimate partner violence, just divorced, marital conflict, marital discord, marital issues, marital problems, marital separation, marital strain, marital stress, marriage ended, marriage not working, of unhealthy relationship, pain marital separation, partner abuse, partner violence, pending divorce, physical abuse domestic, poor family relationship, recent marital separation, recently divorced, relationship breakdown, relationship ended, relationship issues, relationship violence, remaried, remarriage, remarried, remarry, remarrying, sexual trauma interpersonal, the marital separation, to interpersonal violence, trauma domestic, trauma interpersonal violence, trial separation, unhealthy relationship, violence current partner, violence domestic, violence in family, violence intimate partner, violence past |

| | | |
|---|---|---|
| | | partner, violence relationship, violence relationship abusive, violence towards ex, widowed, widower, wife left home, xwife |
| Military Sexual Trauma | 11 | military sex trauma, military sexual abuse, military sexual assault, military sexual harassment, military sexual trama, military sexual trauma, mst, placed military sexual, rape military, sexual assault military, trauma military sexual |
| Justice and Law | 299 | aggravated assault on, ago court ordered, any legal charges, appear in court, aquitted, arraignment, arressted, arrested by police, arrhested, arson, assault charge, at correctional facility, at federal prison, at the prison, attend court, awaiting charges, bailed him out, buglary, bugulary, bulglary, burgalry, burgarly, burgerly, burglaries, burglarized, burglarly, burglary, burglery, burgulary, burgurlary, burlary, carjacking, cellmate, charged with crime, charges court, charges jail time, charges trial, child support which, conditional probation, continue court, convicted, convictiion, conviction, convictions, correctional facilities, correctional facility for, counterfeiting, court appearances, court cases, court dates, court hearing, court mandated, court note, court on, court ordered, court outreach, court proceeding, court referred, courtdate, courtmartial, criminal charge, criminal charges, current charges court, department of corrections, domestic violence case, domestic violence charges, driving violation, embezzelment, embezzlement, encarcerated, encarceration, engaged in illegal, exhonorated, exonerated, extradited, extradition, facilities prison, federal court, federal prison in, fellony, felon, felonies, felonious, felonius, felonly, felony, felonys, first court hearing, for public intoxication, forgery, from jail on, go to prison, has court, have pending legal, his upcoming court, house arrest, illegal activities, |

| | | |
|---|---|---|
| | | illegally, imprisioned, imprisionment, imprison, imprisoned, imprisonment, imprisonments, in correctional facility, in court, in jail, in prison, incacerated, incaceration, incacerations, incaracerated, incaraceration, incaration, incarcarated, incarcaration, incarcerate, incarcerated, incarceratedfor, incarceratin, incarcerating, incarceration, incarcerations, incarcerrated, incarcerted, incarcertion, incarcination, incarcirated, incarciration, incarcirations, incarcration, incarecerated, incarerated, incareration, incarserated, incrcerated, indictment, indictments, injail, inmate, inmates, inprisoned, issues probation, jail for and, jail for domestic, jail for drugs, jail for it, jail for public, jail inmate, jail issued clothing, jail on bond, jail on his, jail or legal, jail or prison, jail prison time, jailed, jailings, jailtime, justice involvement, justice outreach, justice program note, kidnapping, larcency, larcenies, larceny, law enforcement, lawsuit, lawsuits, legal charges, legal difficulties, legal involvement, legal issues, legal problems, legal situation, legalproblems, lifetime sex offender, manslaughter, marshalled, midemeanor, misdameanor, misdeameanor, misdeamenor, misdeamnor, misdeamnors, misdeamor, misdeamors, misdeanor, misdemanor, misdemeaner, misdemeaners, misdemeanor, misdemeanors, misdemeanour, misdemenor, misdemenors, misdomeanor, missed court, missed court hearing, note court hearing, occupation prison, of assault charge, on probation specify, onparole, onprobation, or federal prison, or prison for, others incarceration, parol, parold, parole specify offense, parole upcoming court, paroled, paroles, paroles registered sex, pending court appearances, penetentiary, penitentary, penitentiaries, penitentiary, penitentury, pennitentiary, |

| | | |
|---|---|---|
| | | police custody, porbation, prison as, prison at, prison due, prison due to, prison financial problems, prison first, prison for, prison he, prison he was, prison in, prison inmates, prison jail total, prison juvenile, prison juvenile detention, prison now, prison record, prison sentence, prison sentence, prison time, prison time was, prison until, prison went, prison where, prison where he, prison yes, prison yes financial, probation, probation and possession, probationary, probations, problems legal, prosecuted, prosecution, prosecutor, public intoxication, rearrested, recent arrest, recent court, registered sex offender, reincarcerated, released from prison, released on bail, robbed, robberies, robbery, robbing, roberry, robery, service court, several arrests, sex offender registry, shoplifing, shoplifiting, shoplifted, shopliftin, shoplifting, subpoenaed, sued, the federal court, the legal situation, their incarceration, thievery, to prison for, to prison he, to prison in, treaspassing, trepassing, upcoming court, upcoming court hearing, vandalism, veteran justice outreach, veterans justice outreach, violating his probation, violence case, violence charges, went to court, yes prison |
| Job Instability | 156 | and getting job, backrupcy, backruptcy, banckruptcy, bancruptcy, bankrupt, bankrupted, bankruptsy, bankrutcy, banruptcy, brief employment, concerning employment, consult employment, current employment none, difficulty maintaining employment, dismissed from job, employment agencies, employment application, employment applications, employment assistance, employment awaiting, employment desired, employment discharge, employment issues, employment loss, employment no, employment none, employment problems, employment program, |

| | | |
|---|---|---|
| | | employment soon, employment termination, financial employment issues, finding employment, finding job, fired recently, for job search, getting job, have no job, incomeless, job application, job apply, job coach or, job discharge, job ended, job fair, job financial pain, job interview assisted, job loss, job losses, job no, job out, job placement, job search, job seeking, job soon, jobeless, jobless, joblessness, laid off, lay offs, layoffs, locating employment, looking for employment, lose her job, lose job, loss of employment, loss of job, lost her job, lost his employment, lost his job, lost his job, lost job, marginal employment, nemployed, no employment, no income coming, none employment, nonemployed, obtaining employment, of finding job, of job search, potential employment, quit her job, recent job loss, rehab employment application, rehabilitation employment, resume employment, seeking employment, submit employment applications, supportive employment program, to job interview, to job loss, to lose job, uemployed, uemployment, uenemployment, uenmployed, umemployable, umemployed, umemployment, umeployed, umployed, underemployed, unemloyed, unemmployed, unemmployment, unemploed, unemploeyed, unemploid, unemploued, unemploy, unemployability, unemployabilty, unemployable, unemployablity, unemployd, unemploye, unemployed, unemployee, unemployeed, unemployement, unemployemnt, unemployemtn, unemployent, unemployer, unemployes, unemploymed, unemployment, unemploymnet, unemply, unemplyed, unemplyment, unempoloyed, unempolyed, unempolyment, unempoyed, unempoyment, unempployed, unenployed, uneployed, uneployment, unepmloyed, unmeployed, |

| | | |
|---|---|---|
| | | unmployed, unmployment, unpayed, unployed, very stressful job, voc rehab, vocational assistance, vocational rehab, vocational rehabilitation, with job placement, with job resources, worker compensation, yes brief employment |
| Food Insecurity | 41 | apply for food, applying for food, assistance program snap, availability food planning, benefits food, cash benefits food, child support snap, difficulty obtaining food, eligible for food, emergency food, food access concern, food bank, food banks, food insecurities, food insecurity, food instability, food pantry, food security none, food shortage, food stamp, food stamps, food supply allowing, general relief food, homelessness food, insufficient food supply, meals on wheels, no food access, or snap, public assistance food, receiving meals, relief food, snap based, snap office, snap referral, stamps benefits other, stamps food, stamps general relief, stamps temporary assistance, stressors food, support snap food, to food safety |
| Housing Instability | 253 | afc, aliviane, altamont, alternate housing, americorps, amha, armory, assistance with housing, at bridge housing, ballington, benilde, boudicca, bridgehouse, bristlecone, caap, caaps, cabrillo, campground, carrfour, ccss, ceda, cfdfl, chep, cloudbreak, cmha, cmty, community housing corp, community housing with, concerned about housing, concerns about housing, couch surfing, crc, crosspoint, crrc, cvaf, dcha, dchv, decision regarding housing, degeorge, deim, demontford, dhs, dmha, dom, domicilary, domiciliaries, domiciliary, domicillary, domicilliary, doms, dorm, dormitory, dorms, drrtp, drvrp, dshs, econolodge, enphront, evicted, evicting, eviction, eviction notice, exodus, fema, financial housing, for housing placement, forclosure, foreclose, foreclosed, foreclosure, gdp, get into housing, gpd, |

| | | |
|---|---|---|
| | | gpd program, grace mary manor, guesthouse, hacola, half way house, halfway house, hchv, hcmi, hcrv, hdom, hftb, hiot, hmis, home lost, home placement, homeles, homeless, homelessnes, homelessness, homelesss, homelss, homesless, homestead, homless, hopwa, hostel, housed, housing assistance, housing authority, housing challenges, housing concerns, housing coordinator, housing financial, housing issues, housing lack, housing obtaining, housing placement, housing plan, housing problems, housing program, housing programs, housing referred, housing temporarily, housing unstable, hud, hud vash, huds, hudvash, hvaf, hvehf, hvp, hvrp, hvse, hvsep, hwvp, inadequate housing, inn, issues current living, jsi, kcha, kcvp, lack of housing, legal housing, liberty house, lmha, lost his home, mcvet, mcvets, mdha, mdic, mhicm, mhrrtp, michm, microtel, montachusett, motel, motels, nazcare, nbth, neshv, nmvic, no housing, notice to vacate, nwpp, nycha, obtaining housing, odyssey house, osac, other problems housing, ozanam, pan handling, pcha, pending legal housing, pennyroyal, per housing authority, phlag, php, plowshares, problems housing, prrc, prrp, prrtp, qhouse, railton, rapid rehousing, rcf, rcfs, rebos, rehoused, resettlement, residences, resources living arrangements, road home, rough living, rrha, sadh, safe housing challenges, sahl, sarp, seeking housing, sharehouse, shelter, shelters, shelther, sheltor, sheltors, shleter, sjvv, sorcc, squatter, squatters, squatting, sro, ssd, ssdi, ssfv, ssvf, street living, sud, tbra, temporary housing, the community housing, the housing authority, tlp, transitional housing, transitional residence, travelodge, tses, undomiciled, unsheltered, unstable housing, usvet, |

| | | |
|---|---|---|
| | | usvets, va supportive housing, vacate, vacated, vacating, vadh, vash, vced, vetbridge, vetsville, vhpd, voc, vop, votr, vrq, vsh, vtc, vtp, vvlp, vvrc, vvsd, wayside, wdva, westcare, wvvh, wwha, xroads, ywca |

Final lexicon concepts per life event used for extraction from unstructured clinical text.
